# Hypothesis-generating analysis of mutational signatures in childhood B-cell acute lymphoblastic leukemia in relation to socio-demographic, genetic, and environmental factors: A report from the Children’s Oncology Group

**DOI:** 10.1101/2025.10.02.25336752

**Authors:** Cassandra J. Clark, Zhanni Lu, Nathan Anderson, Yueqi Li, Ji Eun Park, Michael Love, Erin Marcotte, Adam J. de Smith, Logan G. Spector

## Abstract

**Background:** Signatures of mutational processes (mutSig) have the potential to fingerprint exposures present during tumor development. While several studies have described prevalence and burden of mutSig in pediatric populations, there have been few efforts to date to associate mutational signatures with exposures or lifestyle factors related to cancer risk. Studying relationships between mutSig and pediatric cancer risk factors can inform future etiologic studies and elucidate the critical exposure pathways underlying cancer risk.

**Methods:** Our study population includes n=1,491 B-cell acute lymphoblastic leukemia (ALL) cases from the Molecular Profiling to Predict Responses to Therapy (MP2PRT) and a subset of n=856 overlapping cases enrolled in the Childhood Cancer Research Network (CCRN), diagnosed with first primary ALL at ages 0-22 years. We estimated associations between mutSig and demographic and socio-economic factors as Bayesian point estimates and 80% credible intervals (presented as PE [80% CI]) using the *Diffsig* model. The multivariable models included sex, age at diagnosis, either reported race and ethnicity or derived inferred genetic ancestry, cytogenetic subtype, ALL polygenic risk score, parental age at case birth, small-area socio-economic status (SES), and latitude; the latter three variables were available only for the n=856 cases overlapping with CCRN.

**Results:** SBS2 and SBS13 (related to *APOBEC* mutagenesis) were strongly associated with patient sex, reported race and ethnicity, genetic ancestry, and area-level SES. Male cases had strongly decreased relative burdens of SBS2 (−0.26 [−0.41 to −0.10]) and SBS13 (−0.38 [−0.55 to −0.23]) as compared to females. Hispanic/Latine cases had significant enrichment of SBS2 (0.36 [0.22-0.52]) and SBS13 (0.45 [0.29-0.60]) as compared to non-Hispanic Whites; inferred Admixed American ancestry was also associated with enrichment of SBS2 and SBS13 (0.69 [0.50-0.89], 0.93 [0.73-1.12], respectively). Clock-like signatures appeared related to both patient and parental age; SBS5 enrichment was associated with increasing SES and older parental age, and suspected clock-like signature SBS8 was enriched in older diagnostic age groups.

**Conclusions:** In this exploratory analysis of mutSig in pediatric B-cell ALL cases, we identified multiple associations between socio-demographic and genetic factors and mutSig. Signatures related to APOBEC activity were strongly associated with sex, pointing to differences in endogenous immune function between male and female ALL cases. Clock-like signatures show associations with parental and patient age as well as SES, potentially indicating some age-dependent differences. These results should inform future etiologic studies and hypothesis generation for those signatures with presently unknown etiologies.

## I. INTRODUCTION

Childhood acute lymphoblastic leukemia (ALL) is a hematological malignancy that arises from immature B- and T-lymphoid cells.^1^ While long-term survival for ALL exceeds 90%, survivors have an increased risk of adverse outcomes later in life, including chronic illnesses and second malignancies.^2–5^ Despite a decrease in the incidence of cancer overall in the United States, the incidence of childhood ALL has continued to increase, particularly among children of Hispanic ethnicity,^6,7^ underscoring the importance of primary prevention.

The etiology of ALL is likely multifactorial and attributable to both environmental exposures and genetic susceptibility.^1^ The development of preleukemic cells commonly occurs after an initiating genetic mutation *in utero*, with an additional genetic event required for overt ALL to manifest.^8–12^ To date, ALL has been associated with several exogenous exposures such as maternal and child infections,^13–17^ air pollution,^18–23^ pesticides,^24–27^ and benzene,^28–32^ but the ability of traditional population-based epidemiologic studies to identify precise etiologic agents or characterize exposure pathways underlying observed associations is limited. The processes that produce the driver mutations that strongly influence cancer presentation and outcomes are accompanied by signatures of mutational processes.^33,34^ While mutational signatures (mutSig) mostly do not directly affect disease progression and outcomes, they may represent a fingerprint of the insults that were present during mutagenesis. Thus, studying relationships between mutSig and cancer can inform future etiologic studies and elucidate the critical exposure pathways underlying cancer risk.

MutSig can be classified based upon standards established in the Catalogue of Somatic Mutations in Cancer (COSMIC) Mutational Signatures: single (SBS) and doublet (DBS) base substitutions, indels (ID), copy number (CN), structural variants (SV), and RNA single base substitutions (RNA SBS).^35^ A proportion of these signatures have been assigned a proposed etiology based upon a combination of observational and experimental data in humans, animals, or cell lines.^36,37^ Of the documented mutSig to date, roughly a third of SBS and half of IDs have not been assigned a putative etiology.

Pediatric cancers generally have fewer mutations than adult cancers, and mutational burden is strongly correlated with age at diagnosis.^34,38,39^ Our current understanding of the epidemiology of mutational signatures in pediatric cancers is very limited. To date, two pediatric pan-cancer studies have been conducted to describe the broad prevalence of these signatures, one in North America (NA, 27 cancer subtypes) and the other in Europe (EU, 24 cancer subtypes).^38,39^ Of the thirty SBS signatures that have been well-defined in adult cancers,^34^ 29 were observed in the NA cohort and 17 in the EU cohort. Common signatures included those associated with DNA replication and repair defects^40^ and clock-like signatures that tend to be associated with age at diagnosis.^41^ Compared to other pediatric cancers in these cohorts, hematologic malignancies, including ALL, tended to have low to moderate mutational burdens.

While these studies described the prevalence and burden of mutSig among pediatric populations,^38,39^ there have been few efforts to date to associate mutational signatures with exposures or lifestyle factors related to cancer risk. Perhaps the most well-described in children is the enrichment of UV radiation-related mutSig in aneuploid subtypes of pediatric B-ALL.^42^ This finding prompted hypothesis generation on the topic, including a parallel epidemiologic study finding that UV exposure is associated with increased ALL risk.^43^

There remains an opportunity to examine the relationships between mutSig, pediatric cancer, and socio-demographic and environmental characteristics, including those that may be related to cancer risk. As childhood cancers exhibit fewer mutations per megabase than adult cancers, the less-mutated environment of pediatric tumors may facilitate more expeditious identification of these associations. The goal of this hypothesis-generating analysis is to evaluate potential associations between mutSig and socio-demographic and environmental factors in a population of children with B-ALL, leveraging multiple sources of clinical and demographic data.^44,45^

## II. METHODS

### II.a. Study Population and Data Sources

This analysis leverages data from the Molecular Profiling to Predict Responses to Therapy (MP2PRT) study of B-ALL (dbGaP accession number phs002005.v1.p1) and the Childhood Cancer Research Network (CCRN; ACCRN07). Both projects utilize data and patients enrolled on protocols from the Children’s Oncology Group (COG), an international clinical trial cooperative of more than 200 hospitals in North America, Canada, Australia, and New Zealand. MP2PRT supports the analysis of biospecimens collected in NCI-sponsored clinical trials and contains whole genome-sequencing (WGS) data for n=1,491 B-ALL cases, from which mutSig have already been called by Chang et al.; full population selection and sample details can be found there.^46^ The cases in this study are predominantly standard-risk B-cell ALL cases enrolled in COG trials, and the population was enriched for relapsed cases. The CCRN registry protocol enrolled cases were diagnosed with first primary ALL at the ages 0 to 22 years between December 2007 and December 2017. The subset of n=856 cases that overlap with MP2PRT have additional data available, including address at the time of diagnosis for each case and contact data from up to two parents or guardians. This protocol has been approved by the University of Minnesota IRB.

We geocoded these addresses for all U.S. participants; addresses that were resolved to an exact address point or street address were matched to their census block, latitude and longitude. Addresses that were only geocoded to a lower resolution (street name, zip code, city, or unmatched) were considered a poor geocoding match and were excluded from analyses utilizing geocode-derived data. The Yost index, a measure of area-based socioeconomic characteristics based on seven metrics from the American Community Survey, was assigned to each census tract and categorized into quintiles.^47^ We linked the CCRN-enrolled parents and guardians to LexisNexis to receive month and year of birth, as guardian or parental age was not collected in the CCRN protocol, to estimate parental age.

### II.b. Mutational Signatures, Genotyping, and Polygenic Risk Scores

Mutational signatures were called by Chang et al.; the methods used are fully described in that publication.^46^ In short, the authors profiled mutational signatures by fitting single nucleotide substitutions per 96 tri-nucleotide contexts and patterns of insertion deletions (indels) to signatures published in COSMIC V3.2 using the MutationalPatterns R package.^48^

Germline genotype data for all MP2PRT samples were generated using the Genome Analysis Toolkit (GATK v4.6.1.0) best practices workflow for germline short variant discovery.^49^ First, a dataset of 1,369,675 single nucleotide polymorphisms (SNPs) equivalent to those included on the Illumina Global Diversity Array (GDA) was generated from the germline WGS BAM files available on the NCI Genomic Data Commons server. SAMtools^50^ was used to filter BAM files for reads overlapping an autosomal marker on the GDA. Genotype data were processed according to best practices, with variants excluded for missingness (>0.05), MAF (<0.01), Hardy-Weinberg equilibrium (p<10^−7^), and missingness differences by sex (p<10^−7^) before imputation on the TOPMED Imputation Server.^51^ Individual genotypes with a posterior probability ≥ 0.85 were retained, and variants with a low R^2^ (<0.5) were removed. Similarly, variants were excluded for missingness (>0.05), MAF (<0.01), Hardy-Weinberg equilibrium (p<10^−7^), and missingness differences by sex (p<10^−7^). In total, 8,202,037 SNPs were retained.

A polygenic risk score (PRS) for childhood ALL was calculated as the sum of effect weights from 17 loci identified in a prior trans-ethnic GWAS meta-analysis.^52^ PRS were standardized as z-scores (x̅ = 1, SD = 1). To reflect the polygenic risk in MP2PRT cases relative to the population, PRS were standardized with an external population of unaffected controls with available genotype data.^14^

### II.c. Genetic ancestry estimation

Global genetic ancestry was estimated from cleaned genotype data using RFMix (v2.03).^53^ RFMix uses a random forest model trained on a selection of 1000 Genomes phase three data (883 African, 535 Admixed American, 621 East Asian, 562 European, 661 South Asian) to produce local ancestry estimates for these superpopulations at windows across the genome for each individual in the dataset. A default window size of 0.2 cM was used. Eight generations since admixture was assumed to best reflect estimates for European and African admixture.^54^ Global ancestry proportions were calculated by summing the proportions of each chromosome assigned to each ancestry group.

### II.d. Statistical Analysis

Associations between mutSigs and socio-demographic, genetic, and environmental factors were evaluated using *Diffsig.*^55^ *Diffsig* is a statistical modeling framework for estimating how one or more risk factors are associated with predefined mutational signatures. This method estimates the association with a hierarchical Bayesian Dirichlet-Multinomial model based on three key pieces of information: predefined set of mutSigs (e.g. COSMIC SBS signatures), risk factors, and mutation counts summarized by COSMIC SBS and ID mutational profiles (e.g. A[C>A]A, A[C>A]C,…,T[T>G]T) for all samples. For each variable (risk factors), a univariate Diffsig model was run with the COSMIC V3 SBS mutational signatures. We limited our analysis to those signatures comprising at least 1% of the total mutational burden on average (i.e., with a contribution of at least 1%) in the entire MP2PRT population. *Diffsig* provided Bayesian point estimates and credible intervals for the relative association between each variable and mutSigs.

For each model, the dependent variable was either the SBS or ID signatures. The independent variables (those marked with an asterisk* are only available in the CCRN overlap population), our *a priori* variables of interest, included sex (reference group: female sex), age at diagnosis (reference group: 1-4 years), self-reported race and ethnicity (reference group: non-Hispanic White), genetic ancestry (reference group: European ancestry), parental age (<25 years, 25-34 years, 35+ years)*, cytogenetic subtype, Yost index as a measure of small-area socio-economic status (SES; reference group: Quintile 1)*, latitude*, and ALL PRS (either per standard deviation increase or above and below the median). Correlation matrices for all variables are shown in **Supplemental Figure S1**. Of the cytogenetic subtypes, only those with at least n=10 cases were included as a category (BCR::ABL1-like, Down syndrome ALL, DUX4-rearranged, ETV6::RUNX1, ETV6::RUNX1-like, hyperdiploid, PAX5alt, TCF3::PBX1, ZNF384-rearranged); the remainder were considered missing. We conducted separate models for reported race and ethnicity and genetic ancestry. Reported race and ethnicity are sometimes used as proxies for genetic ancestry, as they may be moderately to highly correlated. However, reported race and ethnicity may also provide a proxy for aspects of a racialized experience and exposure to systemic and structural racism.^56,57^ We also considered PRS both continuously and dichotomously (above and below the median among all cases). All analyses were stratified by CCRN overlap, with one set of analyses conducted within the entire MP2PRT population (n=1,491), and a second within only those cases overlapping with CCRN (n=856), who have more specific parental age and location information available due to their linkage with CCRN. Unless otherwise specified, the presented results are from the entire MP2PRT population.

## III. RESULTS

### III.a. Population Characteristics and Mutation Frequencies

The full MP2PRT and CCRN overlap populations were demographically very similar with respect to all examined characteristics, including genomic subtypes of B-ALL (**Table 1**). The distribution of age at diagnosis and subtypes were as expected. The average number of overall mutations per megabase was: (1) For 60 SBSs, 2740.1 in MP2PRT cases and 2904.6 in CCRN overlapping cases; (2) For 18 IDs, 676.2 in MP2PRT cases and 660.2 in CCRN overlapping cases, and the relative contributions varied widely by signature for SBS and ID (**Supplemental Table S1**). Thirty-four SBS (**Supplemental Table S2**) and eleven ID (**Supplemental Table S3**) signatures were detected in at least one sample, but comprised <1% of total mutational burden on average, so were unable to be assessed. Thus, we retained 26 SBS and 7 ID signatures in all subsequent analyses.

**Table 1.**
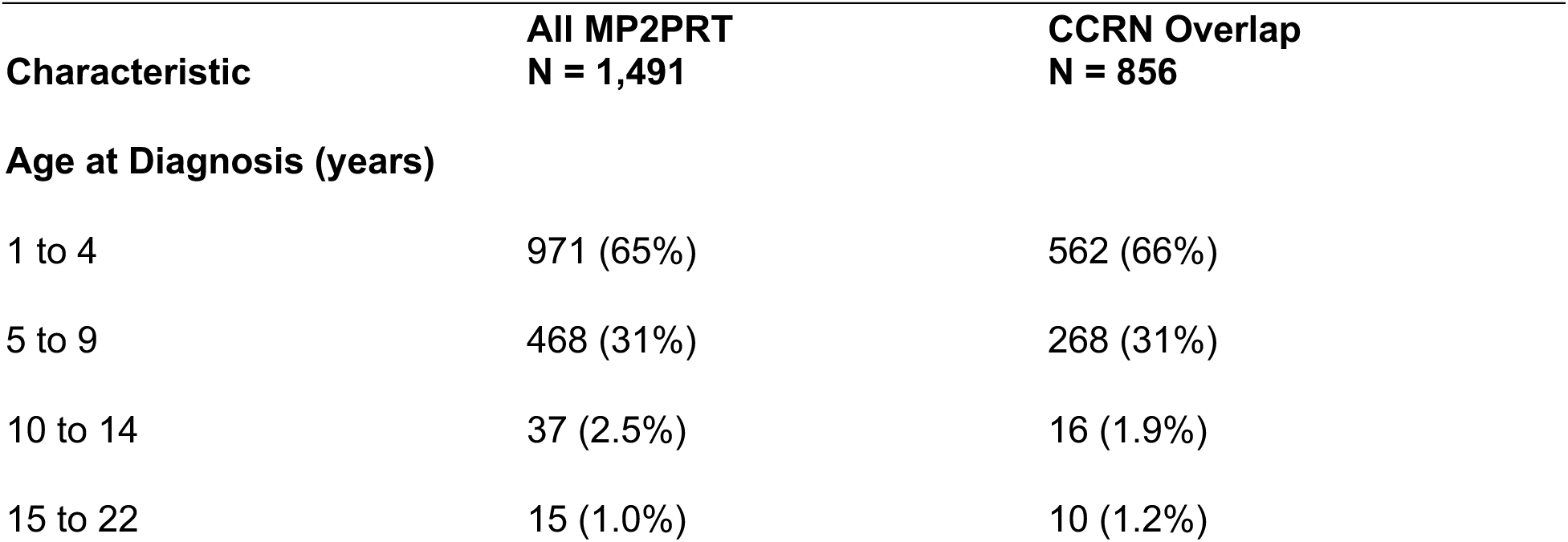
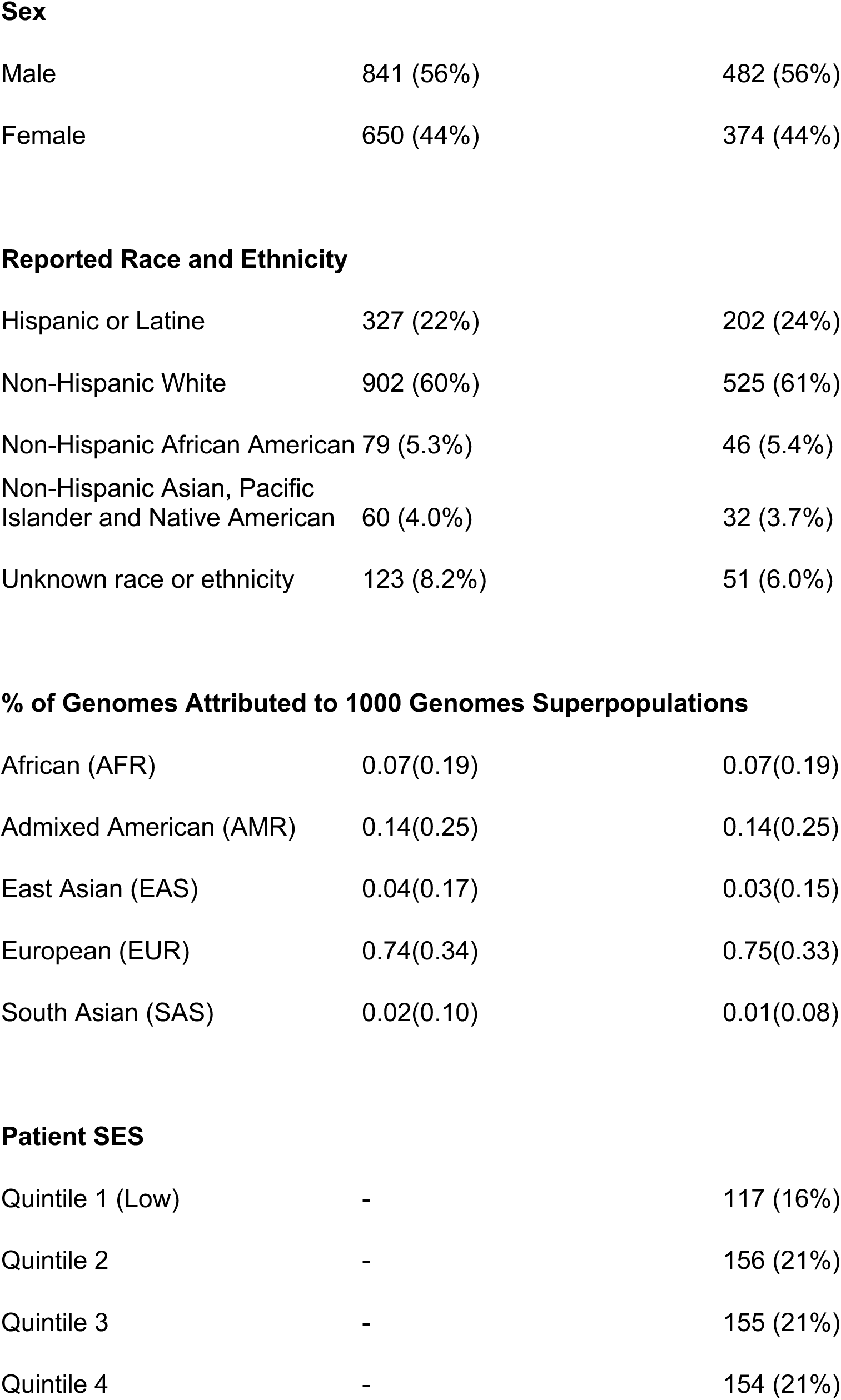
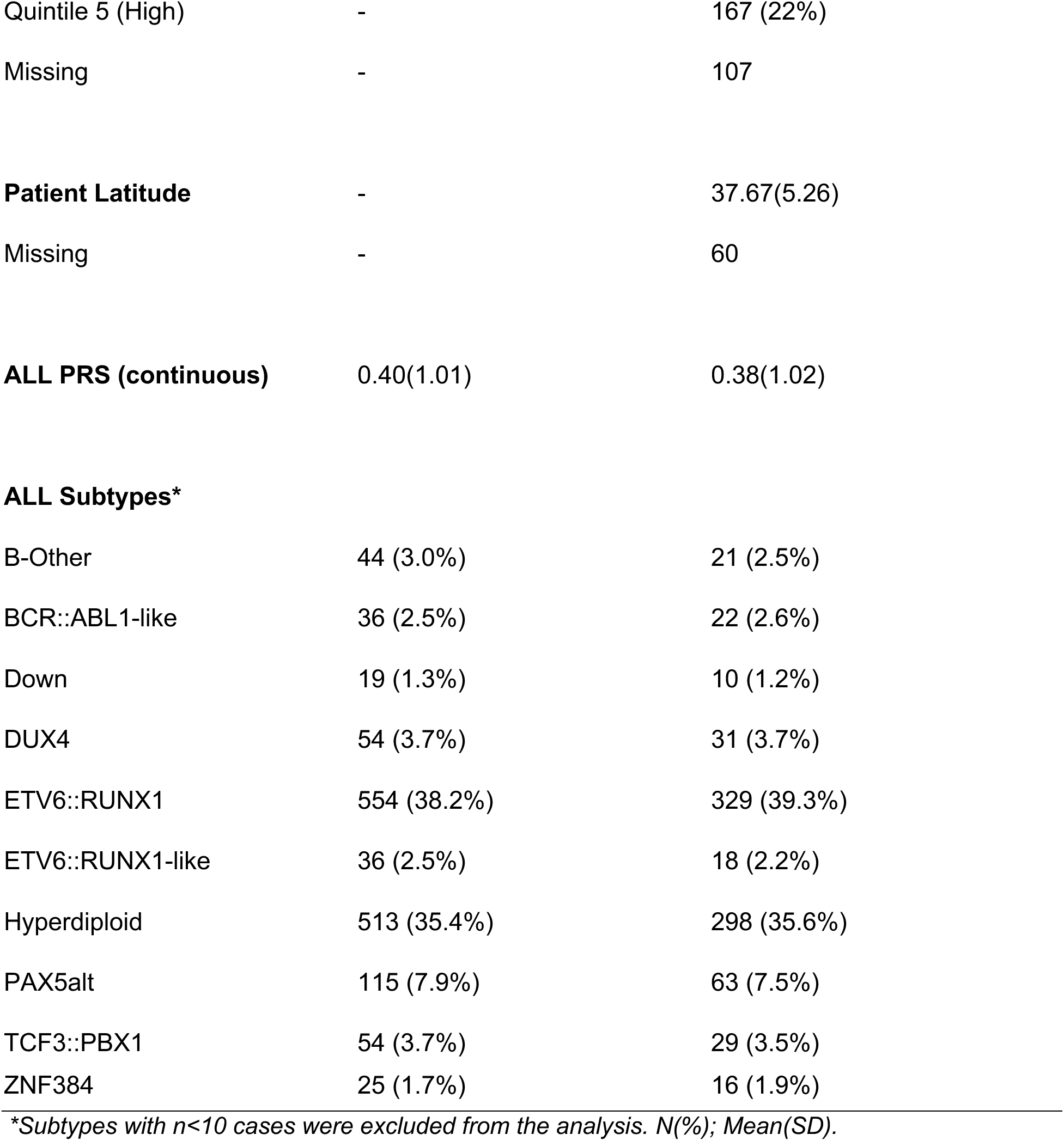
Demographic and Clinical Characteristics of MP2PRT and CCRN cases.

| Characteristic | All MP2PRT<br>N = 1,491 | CCRN Overlap<br>N = 856 |
| --- | --- | --- |
| **Age at Diagnosis (years)** |  |  |
| 1 to 4 | 971 (65%) | 562 (66%) |
| 5 to 9 | 468 (31%) | 268 (31%) |
| 10 to 14 | 37 (2.5%) | 16 (1.9%) |
| 15 to 22 | 15 (1.0%) | 10 (1.2%) |
| **Sex** |  |  |
| Male | 841 (56%) | 482 (56%) |
| Female | 650 (44%) | 374 (44%) |
| **Reported Race and Ethnicity** |  |  |
| Hispanic or Latine | 327 (22%) | 202 (24%) |
| Non-Hispanic White | 902 (60%) | 525 (61%) |
| Non-Hispanic African American | 79 (5.3%) | 46 (5.4%) |
| Non-Hispanic Asian, Pacific<br>Islander and Native American | 60 (4.0%) | 32 (3.7%) |
| Unknown race or ethnicity | 123 (8.2%) | 51 (6.0%) |
| **% of Genomes Attributed to 1000 Genomes Superpopulations** |  |  |
| African (AFR) | 0.07(0.19) | 0.07(0.19) |
| Admixed American (AMR) | 0.14(0.25) | 0.14(0.25) |
| East Asian (EAS) | 0.04(0.17) | 0.03(0.15) |
| European (EUR) | 0.74(0.34) | 0.75(0.33) |
| South Asian (SAS) | 0.02(0.10) | 0.01(0.08) |
| **Patient SES** |  |  |
| Quintile 1 (Low) | - | 117 (16%) |
| Quintile 2 | - | 156 (21%) |
| Quintile 3 | - | 155 (21%) |
| Quintile 4 | - | 154 (21%) |
| Quintile 5 (High) | - | 167 (22%) |
| Missing | - | 107 |
| **Patient Latitude** | - | 37.67(5.26) |
| Missing | - | 60 |
| **ALL PRS (continuous)** | 0.40(1.01) | 0.38(1.02) |
| **ALL Subtypes\*** |  |  |
| B-Other | 44 (3.0%) | 21 (2.5%) |
| BCR::ABL1-like | 36 (2.5%) | 22 (2.6%) |
| Down | 19 (1.3%) | 10 (1.2%) |
| DUX4 | 54 (3.7%) | 31 (3.7%) |
| ETV6::RUNX1 | 554 (38.2%) | 329 (39.3%) |
| ETV6::RUNX1-like | 36 (2.5%) | 18 (2.2%) |
| Hyperdiploid | 513 (35.4%) | 298 (35.6%) |
| PAX5alt | 115 (7.9%) | 63 (7.5%) |
| TCF3::PBX1 | 54 (3.7%) | 29 (3.5%) |
| ZNF384 | 25 (1.7%) | 16 (1.9%) |

*\*Subtypes with n<10 cases were excluded from the analysis. N(%); Mean(SD).*

All results of multivariable models are presented as Bayesian point estimates for the mean effect and 80% credible intervals (presented as PE [80% CI]) for the relative association between each variable and mutSig.

### III.b. Sex

In all models, male B-ALL cases had significantly decreased relative burdens of SBS2 (−0.26 [− 0.41 to −0.10]) and SBS13 (−0.38 [−0.55 to −0.23]) signatures, both related to APOBEC mutagenesis, as compared to female cases (**Figure 1**). Conversely, male cases were enriched in SBS85 (indirect effects of AID, a member of the APOBEC family of enzymes) as compared to female cases (0.16 [0.01-0.32]). Of those signatures with presently unknown etiologies, only SBS19 (unknown) had marginally decreased relative contributions in males as compared to females (−0.16 [−0.32-0.01]).

**Figure 1.**
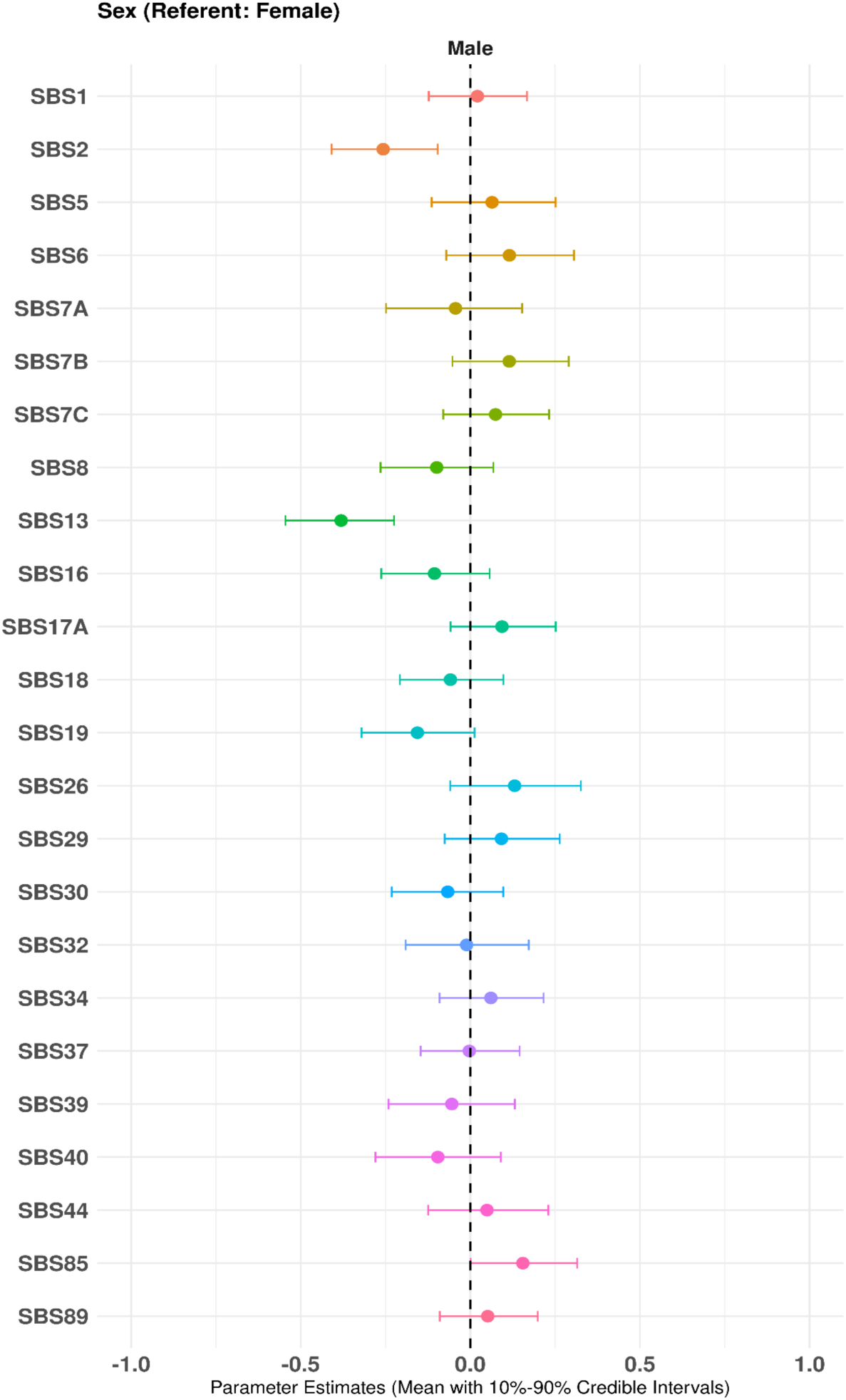
Associations between male sex and relative burden of SBS mutational signatures. Bayesian point estimates represent the mean effect, and error bars represent the 80% credible interval for the relative association between male sex and relative burden of each SBS, as compared to female sex. Adjusted for age at diagnosis, genetic ancestry, cytogenetic subtype, and PRS.

### III.c. Age at diagnosis

Multiple signatures showed age-dependent patterning even after adjustment for other covariates. Age at diagnosis was inconsistently related to SBS2 and SBS13 (APOBEC; **Figure 2**). Of these signatures, only SBS13 had non-significantly decreased relative contributions in the 10-14 (−0.17 [−0.45-0.10]) and 15-22 (−0.29 [−0.67-0.09]) year age groups as compared to the 1-4 year age group. SBS1 (clock-like) was enriched in the 5-9 (0.12 [−0.01-0.26]) and 10-14 (0.15 [−0.02-0.32]) year age groups as compared to the 1-4 year group. Conversely, SBS5 (clock-like) had a decreased contribution to mutational burden in the 5-9 (−0.16 [−0.32-0.02]) and 10-14 (− 0.31 [−0.66-0.05]) year groups and an increased contribution in the 15-22 year group (0.26 [− 0.09-0.62]).

**Figure 2.**
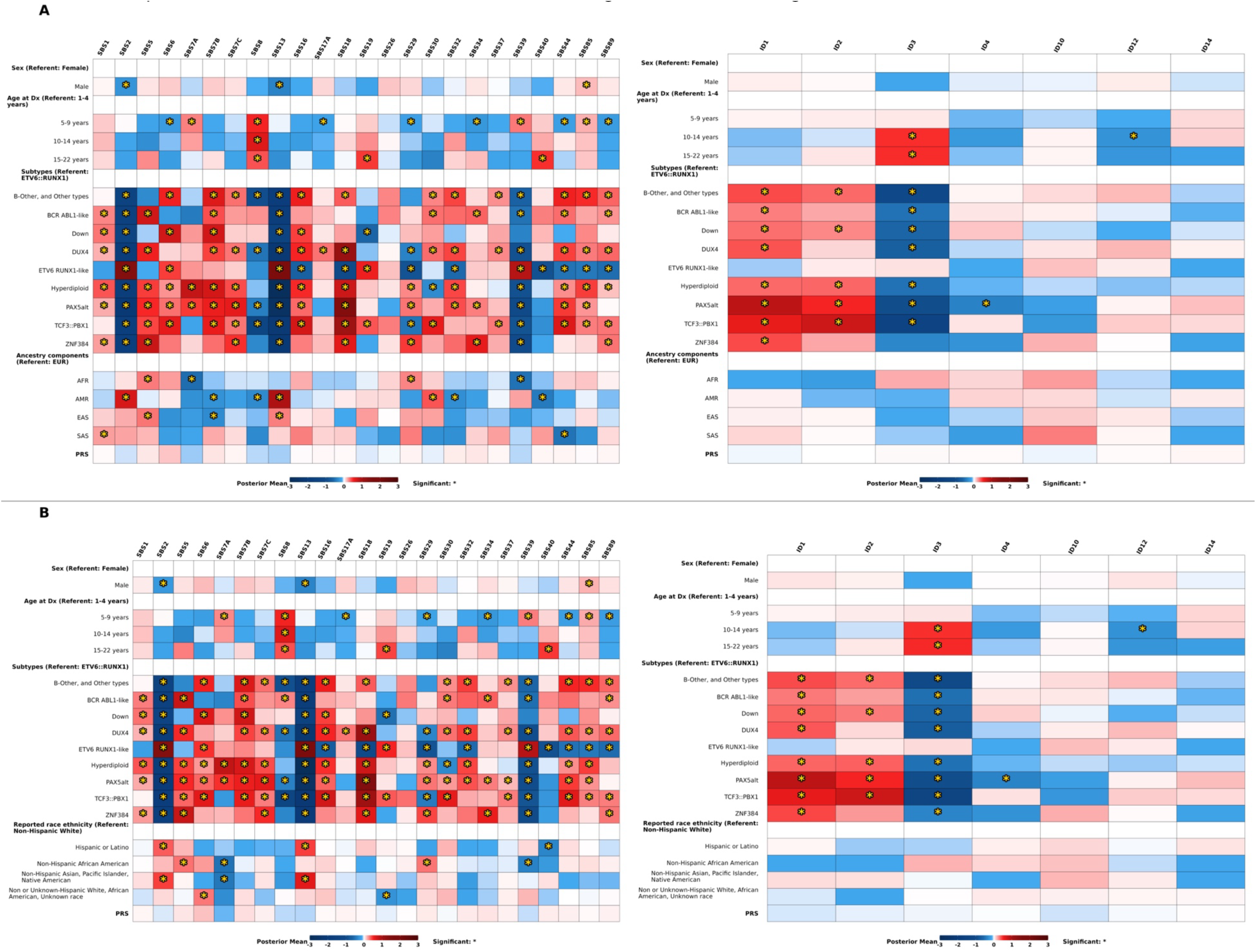
Heatmap of the associations between selected risk factors, SBS, and ID in the MP2PRT population (n=1,491). Panel A) is the multivariable model with ancestry; Panel B) is the multivariable model with reported race and ethnicity. Blue indicates a negative association, while red indicates a positive association; darker colors indicate stronger associations. Significance is denoted with an asterisk.

SBS6 and SBS44 (defective DNA mismatch repair) were decreased in the 5-9 year group (−0.21 [−0.42 to −0.01] and −0.18 [−0.35 to −0.01], respectively) as compared to the 1-4 year group, but did not display other age-dependent patterning. SBS30 (defective DNA repair and replication) appeared inversely associated with increasing age at diagnosis in the 15-22 (−0.39 [−0.84-0.04]) year age group as compared to the 1-4 year group. Interestingly, ID3 (tobacco smoking) was significantly enriched in the 10-14 (0.48 [0.18-0.78]) and 15-22 (0.49 [0.12-0.84]) year age groups as compared to the 1-4 yr group.

Of the signatures with presently unknown etiologies, SBS8 was strongly enriched in all older age groups (5-9 years: 0.52 [0.36-0.68], 10-14 years: 0.65 [0.40-0.90], 15-22 years: 0.39 [0.03-0.75]), as were SBS19 and SBS40. SBS17A (−0.16 [−0.30 to −0.01]), SBS34 (−0.25 [−0.40 to - 0.08]), and SBS89 (−0.21 [−0.34 to −0.07]), also of unknown etiologies, all had decreased contributions in the 5-9 year age group; these associations were generally consistent in the 10-14 year group for SBS34 and SBS89. ID12 (also unknown) was decreased in the 10-14 (−0.27 [− 0.53 to −0.01]) and 15-22 (−0.25 [−0.54-0.05]) year groups as compared to the 1-4 year group.

### III.d. Reported race and ethnicity and genomic ancestry

Reported race and ethnicity, considered separately from inferred genomic ancestry, were strongly associated with multiple signatures even after adjustment for other covariates (**Figure 2**). There were distinct differences in SBS2 and 13 (APOBEC-related signatures) across reported racial and ethnic groups. Hispanic/Latine cases had a significantly higher relative burden of SBS2 (0.36 [0.22-0.52]) and 13 (0.45 [0.29-0.60]) as compared to non-Hispanic White cases. SBS2 (0.45 [0.24-0.66]) and 13 (0.61 [0.39-0.82]) were also strongly enriched in non-Hispanic Asian, Pacific Islander, and Native American or Indigenous cases relative to non-Hispanic Whites.

SBS5 (clock-like) was significantly enriched among non-Hispanic African American cases (0.33 [0.07-0.56]) as compared to non-Hispanic Whites. SBS29 (tobacco chewing; 0.32 [0.09-0.54]) was also enriched among non-Hispanic African American cases as compared to non-Hispanic White cases. SBS7A (UV light exposure; −0.50 [−0.93 to −0.13]) was significantly decreased in non-Hispanic African American and non-Hispanic Asian, Pacific Islander, and Native American or Indigenous cases (−0.48 [−0.93 to −0.08]) relative to non-Hispanic White cases.

Of the signatures with unknown etiologies, the relative burden of SBS8 was decreased among non-Hispanic Asian, Pacific Islander, and Native American or Indigenous cases (−0.23 [−0.50-0.04]) as compared to non-Hispanic White. The relative contribution of SBS40 was also decreased among Hispanic/Latine (−0.36 [−0.57 to −0.15]) and non-Hispanic African American cases (−0.23 [−0.56-0.09]) as compared to non-Hispanic White cases. SBS39 was decreased among non-Hispanic African American cases only (−0.39 [−0.71 to −0.08]).

Proportion of inferred genomic ancestry was also strongly associated with mutSig burden (**Figure 2**). Cases with high proportions of African ancestry had significantly decreased burden of SBS7A (UV light exposure; −0.55 [−0.97 to −0.15]), SBS39 (unknown; −0.50 [−0.85 to −0.15]), and a marginally decreased burden of ID2 (replication slippage; −0.20 [−0.45-0.04]) as compared to cases with high proportions of European ancestry. African ancestry was also associated with relative enrichment of SBS5 (clock-like; 0.30 [0.02-0.58]) and SBS29 (tobacco chewing; 0.28 [0.02-0.54]) as compared to European ancestry.

Admixed American ancestry was associated with significant and strong enrichment of SBS2 and SBS13 (APOBEC activity; 0.69 [0.50-0.89] and 0.93 [0.73-1.12], respectively), and SBS30 (defective DNA base repair; 0.39 [0.18-0.61]). SBS7A (−0.29 [−0.62-0.04]), 7B (−0.28 [−0.52 to - 0.04]), and 7C (−0.18 [−0.37-0.02]; all UV light exposure) were all marginally to significantly decreased in children with high proportions of Admixed American ancestry as compared to European ancestry. SBS8 (unknown clock-like; −0.33 [−0.56 to −0.10]) and SBS40 (unknown; - 0.44 [−0.74 to −0.16]) were inversely associated with increasing Admixed American ancestry.

Enrichment of SBS13 (0.29 [0.05-0.54]) and, to a lesser extent, SBS2 (0.19 [−0.06-0.42]; both APOBEC activity), was associated with increasing inferred East Asian ancestry. Inferred South Asians ancestry was associated with enrichment of SBS1 (clock-like; 0.22 [0.01-0.43]), SBS37 (unknown; 0.22 [−0.01-0.44]), and ID10 (unknown; 0.30 [−0.02 to 0.62]) as compared to increasing European ancestry. Conversely, the relative contributions of SBS44 (defective DNA mismatch repair; −0.49 [−0.97 to −0.01]) were inversely associated with increasing proportion of South Asian ancestry.

### III.e. Polygenic risk score

Increasing ALL PRS was associated with a small number of signatures in multivariate models (**Figure 2**). For each standard deviation increase in PRS, SBS6 (DNA mismatch repair; 0.14 [− 0.01-0.27]) was marginally enriched. When considered dichotomously, SBS6 was significantly enriched in children with PRS higher than the median (0.22 [0.04-0.41]). Conversely, SBS7A was decreased in children with higher PRS scores (−0.20 [−0.40 to −0.01]); SBS30 (defective DNA base excision repair due to NTHL1 mutations; −0.12 [−0.28-0.04]) was also moderately decreased among children with above median PRS scores. No IDs were associated with PRS.

### III.f. Small-area SES

Associations between mutSig and small-area SES were assessed only in the CCRN overlap population (**Figure 3**). In multivariate models, the relative contributions of SBS 2 and 13 (APOBEC) were decreased in Quintiles 3 (−0.11 [−0.33-0.12] and −0.22 [−0.44-0.01], respectively), 4 (−0.12 [−0.34-0.10] and −0.34 [−0.57 to −0.10], respectively), and 5 (−0.19 [−0.41-0.04] and −0.30 [−0.53 to −0.06], respectively) as compared to Quintile 1 (low). SBS5 (clock-like) was consistently enriched in higher quartiles, significantly so in Quartile 5 (0.41 [0.13-0.69]. SBS7a (UV light exposure) was inconsistently associated with SES, being significantly decreased in only Quartile 2 as compared to Quartile 1 (−0.48 [−0.84 to −0.12]. SBS32 (Azathioprine treatment) had consistently significantly decreased relative contribution in Quartiles 2, 3, 4, and 5 as compared to Quartile 1. None of the IDs evaluated appeared to be associated with small-area SES.

**Figure 3.**
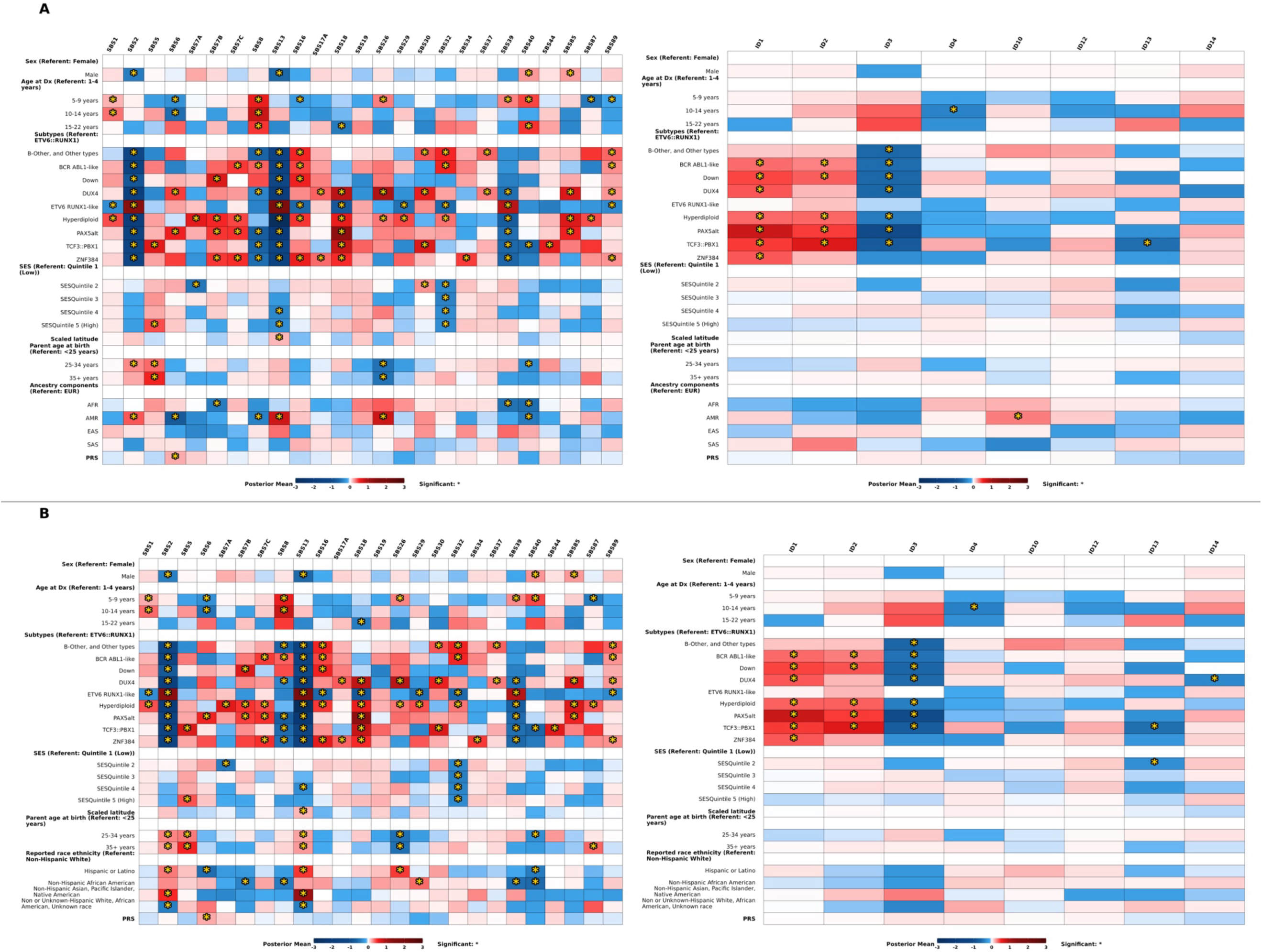
Heatmap of the associations between selected risk factors, SBS, and ID in the overlapping CCRN population (n=856). Panel A) is the multivariable model with ancestry; Panel B) is the multivariable model with reported race and ethnicity. Blue indicates a negative association, while red indicates a positive association; darker colors indicate stronger associations. Significance is indicated with an asterisk.

### III.g. Parental age

After adjusting for other covariates, increasing parental age, both ages 25-34 and 35+ years, was associated with enrichment of SBS2 (APOBEC activity; 0.27 [0.09-0.45] and 0.20 [−0.02-0.42], respectively) and SBS5 (clock-like; 0.34 [0.09-0.62] and 0.53 [0.25-0.81], respectively) as compared to parental ages <25 years (**Figure 3**). SBS13 (APOBEC) was also marginally enriched in the 25-34 year parental group (0.15 [−0.02-0.33]. SBS26 (defective DNA mismatch and repair; 25-34 year group: −0.52 [−0.77 to −0.28], 35+ year group: −0.49 [−0.79 to −0.19]) and SBS40 (unknown; 25-34 year group: −0.30 [−0.53 to −0.06], 35+ year group: −0.23 [−0.50-0.05]) were also marginally to significantly decreased in these groups as compared to children with younger parents. SBS44 (defective DNA mismatch repair) was also somewhat enriched in children with parents in the 25-34 year age group [0.24 [−0.01-0.49].

### III.h. Latitude

Interestingly, increasing latitude had unexpected associations with a few signatures, even after adjusting for other covariates (**Figure 3**). Enrichment of both SBS2 and SBS13 (APOBEC activity; 0.13 [−0.01-0.27] and 0.16 [0.01-0.31], respectively) was associated with increasing latitude. Enrichment of SBS19 (unknown) was also associated with increasing latitude, though the confidence intervals were wide (0.13 [−0.03-0.28]. UV light signatures 7A, 7B, and 7C did not appear to be associated with latitude in our models.

## IV. DISCUSSION

In this hypothesis-generating analysis of a population of pediatric B-cell acute lymphoblastic leukemia patients, we identified multiple associations between socio-demographic and genetic factors and mutSig. APOBEC-related signatures SBS2 and SBS13 appear to be those most sensitive to values of the variables under study, and these were strongly associated with patient sex, reported race and ethnicity, and genetic ancestry. APOBEC signatures were also inversely associated with increasing small-area SES. Clock-like signature SBS5 was associated with increasing SES and older parental age, and suspected clock-like signature SBS8 was enriched in older age groups.

SBS2 and SBS13, both associated with APOBEC activity,^58^ were also associated with multiple known ALL risk factors, mainly sex, self-reported race and ethnicity, and ancestry. We also observed the expected enrichment of APOBEC signatures in *ETV6::RUNX1* and *ETV6::RUNX1*-like subtypes. The Apolipoprotein B mRNA Editing Catalytic Polypeptide-like (APOBEC) family of cytidine deaminases have broad functions generally related to immunity. Activation-induced cytidine deaminase (AID) deaminates cytidines in the immunoglobulin variable gene region during class switch recombination and increases antibody-antigen affinity through somatic hypermutation.^59–62^ Certain APOBEC enzymes - including those in the APOBEC3 family that are thought to underlie SBS2 and SBS13 signatures^63^ - also have limited anti-viral function.^64^ APOBEC family enzymes likely contribute to carcinogenesis through DNA mutator activity, and potentially uncontrolled hypermutation. ^65^ Interestingly, however, loss of APOBEC-encoding genes (e.g., *APOBEC3B*) has also been associated with increased cancer risk.^66,67^ Enzymes of the APOBEC family are significant contributors to mutational burden in adult cancers, including lung, head/neck, and breast cancers, among others.^33,68^ The prevalence of these mutations in pediatric cancers is less well-characterized. However, a few studies have compared frequencies in ALL cases and across ALL subtypes. In general, we observed similar prevalence and enrichment of SBS2 and 13, both attributed to activity by the APOBEC family of enzymes, as reported in the literature. Brady et al. observed SBS2 and 13 in 43% of *ETV6::RUNX1* and 100% of *ETV6::RUNX1*-like cases, respectively.^42^ In another study of 689 B-ALL cases, SBS2 and 13 (referred to in this study as T-2 and T-7) were also highly enriched in *ETV6::RUNX1* cases.^69^ SBS2 and 13 were also seen in a subset of high-mutational burden B-ALL cases, comprising ∼5-6% of the total mutational burden, on average.^70^ Reported values do differ somewhat by population, which could be due to regional population variance or to differences in methodology, data processing, and algorithm choice.^71^ For instance, Thatikonda et al. did not observe SBS2 and 13 in ALL cases,^38^ and Gröbner et al. observed only very limited enrichment (<1% of total mutational burden) in the ALL subgroup.^39^ Notably, these subgroups were relatively small (n=22 B-ALL-other cases and n=16 hypodiploid cases; and n=61 non-hypodiploid B-ALL cases and n=20 hypodiploid cases, respectively), and enrichment in our study and in other studies appears strongly subtype-dependent.^42,72^

Enrichment of the APOBEC-related signatures SBS2 and 13 in childhood ALL cases specifically with *ETV6::RUNX1* fusion or the *ETV6::RUNX1*-like phenotype points to possible subtype-specific effects of early-life infections or immune dysregulation. The increased enrichment in APOBEC-related signatures among female cases suggests that endogenous processes may underlie the sex difference in ALL risk. Similar disparities in SBS2 burden have also been seen in adult cases of esophageal cancer (SBS2).^73^ Sex-specific differences in immune response are well-documented, owing to both genes and hormones.^74^ Differences in immune function likely begin *in utero*,^75,76^ as male testes begin producing androgens (which suppress immune cell activity)^77^ at around ten weeks gestation.^78^ Throughout the lifecourse, males generally have a stronger pro-inflammatory response than females.^74,79^ Autoimmune diseases are more common in females,^80–82^ and immunotherapy treatment efficacy and outcomes are notably worse among female patients (among adults).^73,83,84^ Our results, taken together with the literature, point to differences in endogenous immune function between male and female ALL cases.

With respect to environmental or behavioral exposures, several signatures are thought to be associated with tobacco smoking and chewing. We observed some enrichment of SBS29 (tobacco chewing) in cases of non-Hispanic African American reported race and, separately, cases of African ancestry. Additionally, ID3 (tobacco smoking) was significantly enriched in the 10-14 and 15-22 year diagnostic age groups as compared to younger cases. It is unclear what these enrichments may be representing. Epidemiologic case-control studies of maternal tobacco use during gestation have generally yielded null associations, though paternal tobacco use shows some suggestive effects.^85–88^ Early-life tobacco smoke exposure as inferred by neonatal measures of DNA methylation at the *AHRR* gene (a known biomarker of tobacco exposure) has been associated with an increased frequency of somatic gene deletions among ALL cases^89,90^ and an increased frequency of RAG recombination-mediated deletions at diagnosis,^91^ suggesting potential leukemogenic effects, although further research in this area is warranted.

We observed the expected enrichment of UV-related mutational signatures in aneuploid subtypes of childhood B-ALL (primarily hyperdiploidy) as compared to *ETV6::RUNX1* cases. This has been reported elsewhere, with particular enrichment in patients with intrachromosomal amplification of chromosome 21 (iAMP21);^38,42^ intriguingly, in a study of multiply-relapsed ALL patients, the UV-related signatures were found to be acquired at relapse in some patients with hyperdiploidy and iAMP21, suggesting a potential role for UV radiation exposure at different stages of ALL development.^72^ An analysis of mutational timing in iAMP21 cases indicates that UV signatures were acquired after the generation of the iAMP21 chromosome but often before whole chromosome gains.^92^ The UV signatures appeared to precede the clock-like and other signatures of mutagenesis, indicating that they may have been acquired early in development. UV light is capable of penetrating the skin to reach cutaneous blood vessels.^93^ One current hypothesis for the identification of UV-related signatures in ALL and other hematologic malignancies is that preleukemic cells are circulated in peripheral blood,^92,94^ where they may be exposed to UV light and undergo UV-related mutagenesis. Aneuploid preleukemic cells have been seen in both cord and peripheral blood samples from children later diagnosed with ALL.^95,96^ Such correlations seen in case-only analyses may be fingerprints of exposure, but we cannot determine whether they are causal or risk-related. It is not clear from a mechanistic perspective why preleukemic lymphocytes with chromosomal aneuploidies may be more prone to DNA damage by UV exposure, but it does suggest that UV radiation exposure in early life may be a risk factor for childhood ALL. Increasing UV radiation exposure was recently associated with an increased risk of childhood ALL,^43^ and future studies that leverage information on both mutational signatures and UV exposure data among childhood ALL cases are warranted.

We identified a small number of signatures that were associated with polygenic risk of childhood ALL. SBS6 (DNA mismatch repair) seemed to be enriched in children with a high ALL PRS.

Interestingly, SBS7A (UV light exposure) was decreased in children with higher PRS scores, even after adjusting for cytogenetic subtype and other variables. The potential mechanism behind this is unclear. PRS are a quantitative measure of heritable risk due to common variants on an individual basis. Several ALL risk alleles impact genes involved in B-cell development, and may increase disease risk by stalling B-cells at a “vulnerable” phase that is more prone to leukemogenesis.^97^ Thus, a high PRS for ALL could represent having a permissive genetic background for leukemogenesis or genomic instability. Although genomic PRS developed in individuals with mainly European ancestry have been shown to have reduced predictive ability in non-European populations,^98^ the conventional PRS for ALL based on GWAS SNPs and used in this study has been shown to perform similarly across ancestry groups.^99,100^

Our results could inform hypothesis generation for those signatures with presently unknown etiologies, which have been observed in pediatric ALL, AML, and neuroblastoma, among other tumor types.^38,39,42,101^ SBS8, hypothesized to be associated with late replication errors,^40^ was strongly enriched in all older age groups, as were SBS19 and SBS40. SBS8 was also inversely associated with Black race and Hispanic/Latino ethnicity. Our sample size for Asian, Pacific Islander, and Indigenous and Native American children was limited. Nonetheless, we also observed significant enrichment of SBS8 in this group. These patterns were sometimes, though not always, mirrored within the associated inferred ancestry group. SBS40 was also inversely associated with Black race and Hispanic/Latine ethnicity. Efforts to characterize differences between signatures of known and unknown etiologies have revealed that the latter varied in strand bias, G-C content, and nucleotide diversity, leading to hypotheses that some of these signatures may occur outside of gene regions.^37^

This study has several notable limitations. Mutational signatures present many challenges to interpretation due to the rarity of some signatures, the large number of dependent variables (i.e. outcomes), and signatures of unknown etiology. Due to the large number of outcomes tested, it is possible that some of the observed associations arose due to chance. We made efforts to reduce this chance by limiting the number of signatures under consideration by including only those comprising at least 1% of the mutational burden, which cut the number of signatures under consideration by two-thirds. The mutSig results from the MP2PRT B-ALL patients were generated using MutationalPatterns and limited to the fitting of known SNV and indel COSMIC signatures, and thus there may be additional structural variant-related or *de novo* signatures that we were not able to examine. With respect to *Diffsig* and interpreting the results of this model, mutation counts inherently provide information as to the relative abundance of mutational processes for each sample. However, without additional data, it is not possible to determine which risk factors are causally related to the relative appearance of one mutation over another. The estimated associations for each risk factor model are inherently compositional. For example, we may observe positive associations for risk factor *A* and *B* with signature S with coefficients bA > bB > 0, and negative associations with other signatures. A conclusion could be drawn that factor *A* and *B* both have high associations with signature S relative to the other signatures, but it cannot be further said that risk factor *A* has higher association with signature S than factor *B* with signature S.

Our results provide novel data on associations between mutSig and socio-demographic and clinical characteristics among children with ALL. Future epidemiologic studies should explore potential associations between mutSig and geographic and environmental exposures, such as air pollution, which may be related to inflammation and cancer risk. Future studies could also examine associations between mutSig and PRS for other traits of interest, such as birth weight. The results of our exploratory analysis should be used to support future somatically informed case-control or other etiologic studies.

## Data Availability

Data used in the study are available from dbGaP and the Children's Oncology Group.

## ACKNOWLEDGEMENTS

This research is supported by the National Cancer Institute (T32CA099936 and R01CA266105), the Children’s Cancer Research Fund, and grants U10CA180886, U10CA180899, U10CA098543, and U10CA098413. The content is solely the responsibility of the authors and does not necessarily represent the official views of the National Institutes of Health. A.J.D. is a Scholar of Blood Cancer United.

## REFERENCES

1. Pui, C.-H. in Encycl. Cancer (ed. Schwab, M.) 23–26 (Springer, 2011). doi:10.1007/978-3-642-16483-5_57

2. Bansal, N., Blanco, J. G., Sharma, U. C., Pokharel, S., Shisler, S. & Lipshultz, S. E. Cardiovascular diseases in survivors of childhood cancer. Cancer Metastasis Rev. 39, 55–68 (2020).

3. Baytan, B., Aşut, Ç., Çırpan Kantarcıoğlu, A., Sezgin Evim, M. & Güneş, A. M. Health-Related Quality of Life, Depression, Anxiety, and Self-Image in Acute Lymphocytic Leukemia Survivors. Turk. J. Hematol. 33, 326–330 (2016).

4. Hudson, M. M., Mertens, A. C., Yasui, Y., Hobbie, W., Chen, H., Gurney, J. G., Yeazel, M., Recklitis, C. J., Marina, N., Robison, L. R., Oeffinger, K. C., & Childhood Cancer Survivor Study Investigators. Health status of adult long-term survivors of childhood cancer: a report from the Childhood Cancer Survivor Study. JAMA 290, 1583–1592 (2003).

5. Oeffinger, K. C., Mertens, A. C., Sklar, C. A., Kawashima, T., Hudson, M. M., Meadows, A. T., Friedman, D. L., Marina, N., Hobbie, W., Kadan-Lottick, N. S., Schwartz, C. L., Leisenring, W. & Robison, L. L. Chronic Health Conditions in Adult Survivors of Childhood Cancer. N. Engl. J. Med. 355, 1572–1582 (2006).

6. Barrington-Trimis, J. L., Cockburn, M., Metayer, C., Gauderman, W. J., Wiemels, J. & McKean-Cowdin, R. Trends in childhood leukemia incidence over two decades from 1992 to 2013. Int. J. Cancer 140, 1000–1008 (2017).

7. Giddings, B. M., Whitehead, T. P., Metayer, C. & Miller, M. D. Childhood leukemia incidence in California: High and rising in the Hispanic population. Cancer 122, 2867–2875 (2016).

8. Greaves, M. A causal mechanism for childhood acute lymphoblastic leukaemia. Nat. Rev. Cancer 18, 471–484 (2018).

9. Hein, D., Borkhardt, A. & Fischer, U. Insights into the prenatal origin of childhood acute lymphoblastic leukemia. Cancer Metastasis Rev. 39, 161–171 (2020).

10. Marcotte, E. L., Spector, L. G., Mendes-de-Almeida, D. P. & Nelson, H. H. The Prenatal Origin of Childhood Leukemia: Potential Applications for Epidemiology and Newborn Screening. Front. Pediatr. 9, 639479 (2021).

11. Taub, J. W. & Ge, Y. The prenatal origin of childhood acute lymphoblastic leukemia. Leuk. Lymphoma 45, 19–25 (2004).

12. Wiemels, J. L., Cazzaniga, G., Daniotti, M., Eden, O. B., Addison, G. M., Masera, G., Saha, V., Biondi, A. & Greaves, M. F. Prenatal origin of acute lymphoblastic leukaemia in children. Lancet Lond. Engl. 354, 1499–1503 (1999).

13. Maia, R. da R. P. & Wünsch, V. Infection and childhood leukemia: review of evidence. Rev. Saúde Pública 47, 1172–1185 (2013).

14. Geris, J. M., Schleiss, M. R., Hooten, A. J., Langer, E., Hernandez-Alvarado, N., Roesler, M. A., Sample, J., Williams, L. A., Dickens, D. S., Mody, R. J., Ravindranath, Y., Gowans, K. L., Pridgeon, M. G., Spector, L. G. & Nelson, H. H. Evaluation of the Association Between Congenital Cytomegalovirus Infection and Pediatric Acute Lymphoblastic Leukemia. JAMA Netw. Open 6, e2250219 (2023).

15. Francis, S. S., Wallace, A. D., Wendt, G. A., Li, L., Liu, F., Riley, L. W., Kogan, S., Walsh, K. M., de Smith, A. J., Dahl, G. V., Ma, X., Delwart, E., Metayer, C. & Wiemels, J. L. In utero cytomegalovirus infection and development of childhood acute lymphoblastic leukemia. Blood 129, 1680–1684 (2017).

16. Gallant, R. E., Arroyo, K., Metayer, C., Kang, A. Y., de Smith, A. J. & Wiemels, J. L. Associations between early-life and in utero infections and cytomegalovirus-positive acute lymphoblastic leukemia in children. Int. J. Cancer 152, 845–853 (2023).

17. Wiemels, J. L., Wang, R., Zhou, M., Hansen, H., Gallant, R., Jung, J., Mancuso, N., De Smith, A. J., Metayer, C., Kogan, S. C. & Ma, X. Cytomegalovirus proteins, maternal pregnancy cytokines, and their impact on neonatal immune cytokine profiles and acute lymphoblastic leukemogenesis in children. Haematologica 107, 2266–2270 (2022).

18. Filippini, T., Hatch, E. E., Rothman, K. J., Heck, J. E., Park, A. S., Crippa, A., Orsini, N. & Vinceti, M. Association between Outdoor Air Pollution and Childhood Leukemia: A Systematic Review and Dose-Response Meta-Analysis. Environ. Health Perspect. 127, 46002 (2019).

19. Filippini, T., Heck, J. E., Malagoli, C., Del Giovane, C. & Vinceti, M. A review and meta-analysis of outdoor air pollution and risk of childhood leukemia. J. Environ. Sci. Health Part C Environ. Carcinog. Ecotoxicol. Rev. 33, 36–66 (2015).

20. Janitz, A. E., Campbell, J. E., Magzamen, S., Pate, A., Stoner, J. A. & Peck, J. D. Traffic-related air pollution and childhood acute leukemia in Oklahoma. Environ. Res. 148, 102–111 (2016).

21. Heck, J. E., Park, A. S., Qiu, J., Cockburn, M. & Ritz, B. Risk of leukemia in relation to exposure to ambient air toxics in pregnancy and early childhood. Int. J. Hyg. Environ. Health 217, 662–668 (2014).

22. Heck, J. E., Wu, J., Lombardi, C., Qiu, J., Meyers, T. J., Wilhelm, M., Cockburn, M. & Ritz, B. Childhood cancer and traffic-related air pollution exposure in pregnancy and early life. Environ. Health Perspect. 121, 1385–1391 (2013).

23. Ghosh, J. K. C., Heck, J. E., Cockburn, M., Su, J., Jerrett, M. & Ritz, B. Prenatal exposure to traffic-related air pollution and risk of early childhood cancers. Am. J. Epidemiol. 178, 1233–1239 (2013).

24. Bailey, H. D., Infante-Rivard, C., Metayer, C., Clavel, J., Lightfoot, T., Kaatsch, P., Roman, E., Magnani, C., Spector, L. G., Petridou, E., Milne, E., Dockerty, J. D., Miligi, L., Armstrong, B. K., Rudant, J., Fritschi, L., Simpson, J., Zhang, L., Rondelli, R., Baka, M., Orsi, L., Moschovi, M., Kang, A. Y. & Schüz, J. Home pesticide exposures and risk of childhood leukemia: Findings from the Childhood Leukemia International Consortium. Int. J. Cancer J. Int. Cancer 137, 2644–2663 (2015).

25. Bailey, H. D., Fritschi, L., Infante-Rivard, C., Glass, D. C., Miligi, L., Dockerty, J. D., Lightfoot, T., Clavel, J., Roman, E., Spector, L. G., Kaatsch, P., Metayer, C., Magnani, C., Milne, E., Polychronopoulou, S., Simpson, J., Rudant, J., Sidi, V., Rondelli, R., Orsi, L., Kang, A. Y., Petridou, E. & Schüz, J. Parental occupational pesticide exposure and the risk of childhood leukemia in the offspring: findings from the childhood leukemia international consortium. Int. J. Cancer 135, 2157–2172 (2014).

26. Gunier, R. B., Kang, A., Hammond, S. K., Reinier, K., Lea, C. S., Chang, J. S., Does, M., Scelo, G., Kirsch, J., Crouse, V., Cooper, R., Quinlan, P. & Metayer, C. A task-based assessment of parental occupational exposure to pesticides and childhood acute lymphoblastic leukemia. Environ. Res. 156, 57–62 (2017).

27. Karalexi, M. A., Tagkas, C. F., Markozannes, G., Tseretopoulou, X., Hernández, A. F., Schüz, J., Halldorsson, T. I., Psaltopoulou, T., Petridou, E. T., Tzoulaki, I. & Ntzani, E. E. Exposure to pesticides and childhood leukemia risk: A systematic review and meta-analysis. Environ. Pollut. Barking Essex 1987 285, 117376 (2021).

28. Carlos-Wallace, F. M., Zhang, L., Smith, M. T., Rader, G. & Steinmaus, C. Parental, In Utero, and Early-Life Exposure to Benzene and the Risk of Childhood Leukemia: A Meta-Analysis. Am. J. Epidemiol. 183, 1–14 (2016).

29. Heck, J. E., He, D., Contreras, Z. A., Ritz, B., Olsen, J. & Hansen, J. Parental occupational exposure to benzene and the risk of childhood and adolescent acute lymphoblastic leukaemia: a population-based study. Occup. Environ. Med. 76, 527–529 (2019).

30. Houot, J., Marquant, F., Goujon, S., Faure, L., Honoré, C., Roth, M.-H., Hémon, D. & Clavel, J. Residential Proximity to Heavy-Traffic Roads, Benzene Exposure, and Childhood Leukemia-The GEOCAP Study, 2002-2007. Am. J. Epidemiol. 182, 685–693 (2015).

31. Savitz, D. A. & Andrews, K. W. Review of epidemiologic evidence on benzene and lymphatic and hematopoietic cancers. Am. J. Ind. Med. 31, 287–295 (1997).

32. Snyder, R. Leukemia and benzene. Int. J. Environ. Res. Public. Health 9, 2875–2893 (2012).

33. Alexandrov, L. B., Nik-Zainal, S., Wedge, D. C., Aparicio, S. A. J. R., Behjati, S., Biankin, A. V., Bignell, G. R., Bolli, N., Borg, A., Børresen-Dale, A.-L., Boyault, S., Burkhardt, B., Butler, A. P., Caldas, C., Davies, H. R., Desmedt, C., Eils, R., Eyfjörd, J. E., Foekens, J. A., Greaves, M., Hosoda, F., Hutter, B., Ilicic, T., Imbeaud, S., Imielinsk, M., Jäger, N., Jones, D. T. W., Jones, D., Knappskog, S., Kool, M., Lakhani, S. R., López-Otín, C., Martin, S., Munshi, N. C., Nakamura, H., Northcott, P. A., Pajic, M., Papaemmanuil, E., Paradiso, A., Pearson, J. V., Puente, X. S., Raine, K., Ramakrishna, M., Richardson, A. L., Richter, J., Rosenstiel, P., Schlesner, M., Schumacher, T. N., Span, P. N., Teague, J. W., Totoki, Y., Tutt, A. N. J., Valdés-Mas, R., van Buuren, M. M., van ‘t Veer, L., Vincent-Salomon, A., Waddell, N., Yates, L. R., Zucman-Rossi, J., Futreal, P. A., McDermott, U., Lichter, P., Meyerson, M., Grimmond, S. M., Siebert, R., Campo, E., Shibata, T., Pfister, S. M., Campbell, P. J. & Stratton, M. R. Signatures of mutational processes in human cancer. Nature 500, 415–421 (2013).

34. Alexandrov, L. B., Kim, J., Haradhvala, N. J., Huang, M. N., Tian Ng, A. W., Wu, Y., Boot, A., Covington, K. R., Gordenin, D. A., Bergstrom, E. N., Islam, S. M. A., Lopez-Bigas, N., Klimczak, L. J., McPherson, J. R., Morganella, S., Sabarinathan, R., Wheeler, D. A., Mustonen, V., PCAWG Mutational Signatures Working Group, Getz, G., Rozen, S. G., Stratton, M. R., & PCAWG Consortium. The repertoire of mutational signatures in human cancer. Nature 578, 94–101 (2020).

35. COSMIC | Mutational Signatures. at <https://cancer.sanger.ac.uk/signatures/>

36. Zou, X., Owusu, M., Harris, R., Jackson, S. P., Loizou, J. I. & Nik-Zainal, S. Validating the concept of mutational signatures with isogenic cell models. Nat. Commun. 9, 1744 (2018).

37. Hu, X., Xu, Z. & De, S. Characteristics of mutational signatures of unknown etiology. NAR Cancer 2, zcaa026 (2020).

38. Thatikonda, V., Islam, S. M. A., Autry, R. J., Jones, B. C., Gröbner, S. N., Warsow, G., Hutter, B., Huebschmann, D., Fröhling, S., Kool, M., Blattner-Johnson, M., Jones, D. T. W., Alexandrov, L. B., Pfister, S. M. & Jäger, N. Comprehensive analysis of mutational signatures reveals distinct patterns and molecular processes across 27 pediatric cancers. Nat. Cancer 4, 276–289 (2023).

39. Gröbner, S. N., Worst, B. C., Weischenfeldt, J., Buchhalter, I., Kleinheinz, K., Rudneva, V. A., Johann, P. D., Balasubramanian, G. P., Segura-Wang, M., Brabetz, S., Bender, S., Hutter, B., Sturm, D., Pfaff, E., Hübschmann, D., Zipprich, G., Heinold, M., Eils, J., Lawerenz, C., Erkek, S., Lambo, S., Waszak, S., Blattmann, C., Borkhardt, A., Kuhlen, M., Eggert, A., Fulda, S., Gessler, M., Wegert, J., Kappler, R., Baumhoer, D., Burdach, S., Kirschner-Schwabe, R., Kontny, U., Kulozik, A. E., Lohmann, D., Hettmer, S., Eckert, C., Bielack, S., Nathrath, M., Niemeyer, C., Richter, G. H., Schulte, J., Siebert, R., Westermann, F., Molenaar, J. J., Vassal, G., Witt, H., ICGC PedBrain-Seq Project, ICGC MMML-Seq Project, Burkhardt, B., Kratz, C. P., Witt, O., van Tilburg, C. M., Kramm, C. M., Fleischhack, G., Dirksen, U., Rutkowski, S., Frühwald, M., von Hoff, K., Wolf, S., Klingebiel, T., Koscielniak, E., Landgraf, P., Koster, J., Resnick, A. C., Zhang, J., Liu, Y., Zhou, X., Waanders, A. J., Zwijnenburg, D. A., Raman, P., Brors, B., Weber, U. D., Northcott, P. A., Pajtler, K. W., Kool, M., Piro, R. M., Korbel, J. O., Schlesner, M., Eils, R., Jones, D. T. W., Lichter, P., Chavez, L., Zapatka, M. & Pfister, S. M. The landscape of genomic alterations across childhood cancers. Nature 555, 321–327 (2018).

40. Singh, V. K., Rastogi, A., Hu, X., Wang, Y. & De, S. Mutational signature SBS8 predominantly arises due to late replication errors in cancer. Commun. Biol. 3, 421 (2020).

41. Alexandrov, L. B., Jones, P. H., Wedge, D. C., Sale, J. E., Campbell, P. J., Nik-Zainal, S. & Stratton, M. R. Clock-like mutational processes in human somatic cells. Nat. Genet. 47, 1402–1407 (2015).

42. Brady, S. W., Roberts, K. G., Gu, Z., Shi, L., Pounds, S., Pei, D., Cheng, C., Dai, Y., Devidas, M., Qu, C., Hill, A. N., Payne-Turner, D., Ma, X., Iacobucci, I., Baviskar, P., Wei, L., Arunachalam, S., Hagiwara, K., Liu, Y., Flasch, D. A., Liu, Y., Parker, M., Chen, X., Elsayed, A. H., Pathak, O., Li, Y., Fan, Y., Michael, J. R., Rusch, M., Wilkinson, M. R., Foy, S., Hedges, D. J., Newman, S., Zhou, X., Wang, J., Reilly, C., Sioson, E., Rice, S. V., Pastor Loyola, V., Wu, G., Rampersaud, E., Reshmi, S. C., Gastier-Foster, J., Guidry Auvil, J. M., Gesuwan, P., Smith, M. A., Winick, N., Carroll, A. J., Heerema, N. A., Harvey, R. C., Willman, C. L., Larsen, E., Raetz, E. A., Borowitz, M. J., Wood, B. L., Carroll, W. L., Zweidler-McKay, P. A., Rabin, K. R., Mattano, L. A., Maloney, K. W., Winter, S. S., Burke, M. J., Salzer, W., Dunsmore, K. P., Angiolillo, A. L., Crews, K. R., Downing, J. R., Jeha, S., Pui, C.-H., Evans, W. E., Yang, J. J., Relling, M. V., Gerhard, D. S., Loh, M. L., Hunger, S. P., Zhang, J. & Mullighan, C. G. The genomic landscape of pediatric acute lymphoblastic leukemia. Nat. Genet. 54, 1376–1389 (2022).

43. Little, M. P., Mai, J. Z., Fang, M., Chernyavskiy, P., Kennerley, V., Cahoon, E. K., Cockburn, M. G., Kendall, G. M. & Kimlin, M. G. Solar ultraviolet radiation exposure, and incidence of childhood acute lymphocytic leukaemia and non-Hodgkin lymphoma in a US population-based dataset. Br. J. Cancer 130, 1441–1452 (2024).

44. Molecular Profiling to Predict Response to Treatment (MP2PRT) Program | Frederick National Laboratory. at <https://frederick.cancer.gov/node/5949>

45. Jr, M., Lg, S., Md, K., Gh, R., Am, L., Jn, P., Sk, S., Pc, A. & Ja, R. The Children’s Oncology Group Childhood Cancer Research Network (CCRN): case catchment in the United States. Cancer 120, (2014).

46. Chang, T.-C., Chen, W., Qu, C., Cheng, Z., Hedges, D., Elsayed, A., Pounds, S. B., Shago, M., Rabin, K. R., Raetz, E. A., Devidas, M., Cheng, C., Angiolillo, A., Baviskar, P., Borowitz, M., Burke, M. J., Carroll, A., Carroll, W. L., Chen, I.-M., Harvey, R., Heerema, N., Iacobucci, I., Wang, J. R., Jeha, S., Larsen, E., Mattano, L., Maloney, K., Pui, C.-H., Ramirez, N. C., Salzer, W., Willman, C., Winick, N., Wood, B., Hunger, S. P., Wu, G., Mullighan, C. G. & Loh, M. L. Genomic Determinants of Outcome in Acute Lymphoblastic Leukemia. J. Clin. Oncol. 42, 3491–3503 (2024).

47. Yost, K., Perkins, C., Cohen, R., Morris, C. & Wright, W. Socioeconomic status and breast cancer incidence in California for different race/ethnic groups. Cancer Causes Control CCC 12, 703–711 (2001).

48. Blokzijl, F., Janssen, R., van Boxtel, R. & Cuppen, E. MutationalPatterns: comprehensive genome-wide analysis of mutational processes. Genome Med. 10, 33 (2018).

49. Germline short variant discovery (SNPs + Indels). GATK (2024). At <https://gatk.broadinstitute.org/hc/en-us/articles/360035535932-Germline-short-variant-discovery-SNPs-Indels>

50. Danecek, P., Bonfield, J. K., Liddle, J., Marshall, J., Ohan, V., Pollard, M. O., Whitwham, A., Keane, T., McCarthy, S. A., Davies, R. M. & Li, H. Twelve years of SAMtools and BCFtools. GigaScience 10, giab008 (2021).

51. Taliun, D., Harris, D. N., Kessler, M. D., Carlson, J., Szpiech, Z. A., Torres, R., Taliun, S. A. G., Corvelo, A., Gogarten, S. M., Kang, H. M., Pitsillides, A. N., LeFaive, J., Lee, S.-B., Tian, X., Browning, B. L., Das, S., Emde, A.-K., Clarke, W. E., Loesch, D. P., Shetty, A. C., Blackwell, T. W., Smith, A. V., Wong, Q., Liu, X., Conomos, M. P., Bobo, D. M., Aguet, F., Albert, C., Alonso, A., Ardlie, K. G., Arking, D. E., Aslibekyan, S., Auer, P. L., Barnard, J., Barr, R. G., Barwick, L., Becker, L. C., Beer, R. L., Benjamin, E. J., Bielak, L. F., Blangero, J., Boehnke, M., Bowden, D. W., Brody, J. A., Burchard, E. G., Cade, B. E., Casella, J. F., Chalazan, B., Chasman, D. I., Chen, Y.-D. I., Cho, M. H., Choi, S. H., Chung, M. K., Clish, C. B., Correa, A., Curran, J. E., Custer, B., Darbar, D., Daya, M., de Andrade, M., DeMeo, D. L., Dutcher, S. K., Ellinor, P. T., Emery, L. S., Eng, C., Fatkin, D., Fingerlin, T., Forer, L., Fornage, M., Franceschini, N., Fuchsberger, C., Fullerton, S. M., Germer, S., Gladwin, M. T., Gottlieb, D. J., Guo, X., Hall, M. E., He, J., Heard-Costa, N. L., Heckbert, S. R., Irvin, M. R., Johnsen, J. M., Johnson, A. D., Kaplan, R., Kardia, S. L. R., Kelly, T., Kelly, S., Kenny, E. E., Kiel, D. P., Klemmer, R., Konkle, B. A., Kooperberg, C., Köttgen, A., Lange, L. A., Lasky-Su, J., Levy, D., Lin, X., Lin, K.-H., Liu, C., Loos, R. J. F., Garman, L., Gerszten, R., Lubitz, S. A., Lunetta, K. L., Mak, A. C. Y., Manichaikul, A., Manning, A. K., Mathias, R. A., McManus, D. D., McGarvey, S. T., Meigs, J. B., Meyers, D. A., Mikulla, J. L., Minear, M. A., Mitchell, B. D., Mohanty, S., Montasser, M. E., Montgomery, C., Morrison, A. C., Murabito, J. M., Natale, A., Natarajan, P., Nelson, S. C., North, K. E., O’Connell, J. R., Palmer, N. D., Pankratz, N., Peloso, G. M., Peyser, P. A., Pleiness, J., Post, W. S., Psaty, B. M., Rao, D. C., Redline, S., Reiner, A. P., Roden, D., Rotter, J. I., Ruczinski, I., Sarnowski, C., Schoenherr, S., Schwartz, D. A., Seo, J.-S., Seshadri, S., Sheehan, V. A., Sheu, W. H., Shoemaker, M. B., Smith, N. L., Smith, J. A., Sotoodehnia, N., Stilp, A. M., Tang, W., Taylor, K. D., Telen, M., Thornton, T. A., Tracy, R. P., Van Den Berg, D. J., Vasan, R. S., Viaud-Martinez, K. A., Vrieze, S., Weeks, D. E., Weir, B. S., Weiss, S. T., Weng, L.-C., Willer, C. J., Zhang, Y., Zhao, X., Arnett, D. K., Ashley-Koch, A. E., Barnes, K. C., Boerwinkle, E., Gabriel, S., Gibbs, R., Rice, K. M., Rich, S. S., Silverman, E. K., Qasba, P., Gan, W., NHLBI Trans-Omics for Precision Medicine (TOPMed) Consortium, Papanicolaou, G. J., Nickerson, D. A., Browning, S. R., Zody, M. C., Zöllner, S., Wilson, J. G., Cupples, L. A., Laurie, C. C., Jaquish, C. E., Hernandez, R. D., O’Connor, T. D. & Abecasis, G. R. Sequencing of 53,831 diverse genomes from the NHLBI TOPMed Program. Nature 590, 290–299 (2021).

52. Jeon, S., de Smith, A. J., Li, S., Chen, M., Chan, T. F., Muskens, I. S., Morimoto, L. M., DeWan, A. T., Mancuso, N., Metayer, C., Ma, X., Wiemels, J. L. & Chiang, C. W. K. Genome-wide trans-ethnic meta-analysis identifies novel susceptibility loci for childhood acute lymphoblastic leukemia. Leukemia 36, 865–868 (2022).

53. Maples, B. K., Gravel, S., Kenny, E. E. & Bustamante, C. D. RFMix: A Discriminative Modeling Approach for Rapid and Robust Local-Ancestry Inference. Am. J. Hum. Genet. 93, 278–288 (2013).

54. Bryc, K., Durand, E. Y., Macpherson, J. M., Reich, D. & Mountain, J. L. The genetic ancestry of African Americans, Latinos, and European Americans across the United States. Am. J. Hum. Genet. 96, 37–53 (2015).

55. Park, J.-E., Smith, M. A., Van Alsten, S. C., Walens, A., Wu, D., Hoadley, K. A., Troester, M. A. & Love, M. I. Diffsig: Associating Risk Factors With Mutational Signatures. Cancer Epidemiol. Biomarkers Prev. 33, 721–730 (2024).

56. Banaji, M. R., Fiske, S. T. & Massey, D. S. Systemic racism: individuals and interactions, institutions and society. Cogn. Res. Princ. Implic. 6, 82 (2021).

57. Braveman, P. A., Arkin, E., Proctor, D., Kauh, T. & Holm, N. Systemic And Structural Racism: Definitions, Examples, Health Damages, And Approaches To Dismantling. Health Aff. Proj. Hope 41, 171–178 (2022).

58. Chan, K., Roberts, S. A., Klimczak, L. J., Sterling, J. F., Saini, N., Malc, E. P., Kim, J., Kwiatkowski, D. J., Fargo, D. C., Mieczkowski, P. A., Getz, G. & Gordenin, D. A. An APOBEC3A hypermutation signature is distinguishable from the signature of background mutagenesis by APOBEC3B in human cancers. Nat. Genet. 47, 1067–1072 (2015).

59. Salter, J. D., Bennett, R. P. & Smith, H. C. The APOBEC Protein Family: United by Structure, Divergent in Function. Trends Biochem. Sci. 41, 578–594 (2016).

60. Mertz, T. M., Collins, C. D., Dennis, M., Coxon, M. & Roberts, S. A. APOBEC-Induced Mutagenesis in Cancer. Annu. Rev. Genet. 56, 229–252 (2022).

61. Papavasiliou, F. N. & Schatz, D. G. Somatic hypermutation of immunoglobulin genes: merging mechanisms for genetic diversity. Cell 109 Suppl, S35–44 (2002).

62. Suspène, R., Aynaud, M.-M., Guétard, D., Henry, M., Eckhoff, G., Marchio, A., Pineau, P., Dejean, A., Vartanian, J.-P. & Wain-Hobson, S. Somatic hypermutation of human mitochondrial and nuclear DNA by APOBEC3 cytidine deaminases, a pathway for DNA catabolism. Proc. Natl. Acad. Sci. U. S. A. 108, 4858–4863 (2011).

63. Petljak, M., Dananberg, A., Chu, K., Bergstrom, E. N., Striepen, J., von Morgen, P., Chen, Y., Shah, H., Sale, J. E., Alexandrov, L. B., Stratton, M. R. & Maciejowski, J. Mechanisms of APOBEC3 mutagenesis in human cancer cells. Nature 607, 799–807 (2022).

64. Sheehy, A. M., Gaddis, N. C., Choi, J. D. & Malim, M. H. Isolation of a human gene that inhibits HIV-1 infection and is suppressed by the viral Vif protein. Nature 418, 646–650 (2002).

65. Harris, R. S., Petersen-Mahrt, S. K. & Neuberger, M. S. RNA editing enzyme APOBEC1 and some of its homologs can act as DNA mutators. Mol. Cell 10, 1247–1253 (2002).

66. Middlebrooks, C. D., Banday, A. R., Matsuda, K., Udquim, K.-I., Onabajo, O. O., Paquin, A., Figueroa, J. D., Zhu, B., Koutros, S., Kubo, M., Shuin, T., Freedman, N. D., Kogevinas, M., Malats, N., Chanock, S. J., Garcia-Closas, M., Silverman, D. T., Rothman, N. & Prokunina-Olsson, L. Association of germline variants in the APOBEC3 region with cancer risk and enrichment with APOBEC-signature mutations in tumors. Nat. Genet. 48, 1330–1338 (2016).

67. Long, J., Delahanty, R. J., Li, G., Gao, Y.-T., Lu, W., Cai, Q., Xiang, Y.-B., Li, C., Ji, B.-T., Zheng, Y., Ali, S., Shu, X.-O. & Zheng, W. A common deletion in the APOBEC3 genes and breast cancer risk. J. Natl. Cancer Inst. 105, 573–579 (2013).

68. Burns, M. B., Temiz, N. A. & Harris, R. S. Evidence for APOBEC3B mutagenesis in multiple human cancers. Nat. Genet. 45, 977–983 (2013).

69. Ma, X., Liu, Y., Liu, Y., Alexandrov, L. B., Edmonson, M. N., Gawad, C., Zhou, X., Li, Y., Rusch, M. C., Easton, J., Huether, R., Gonzalez-Pena, V., Wilkinson, M. R., Hermida, L. C., Davis, S., Sioson, E., Pounds, S., Cao, X., Ries, R. E., Wang, Z., Chen, X., Dong, L., Diskin, S. J., Smith, M. A., Guidry Auvil, J. M., Meltzer, P. S., Lau, C. C., Perlman, E. J., Maris, J. M., Meshinchi, S., Hunger, S. P., Gerhard, D. S. & Zhang, J. Pan-cancer genome and transcriptome analyses of 1,699 paediatric leukaemias and solid tumours. Nature 555, 371–376 (2018).

70. Newman, S., Nakitandwe, J., Kesserwan, C. A., Azzato, E. M., Wheeler, D. A., Rusch, M., Shurtleff, S., Hedges, D. J., Hamilton, K. V., Foy, S. G., Edmonson, M. N., Thrasher, A., Bahrami, A., Orr, B. A., Klco, J. M., Gu, J., Harrison, L. W., Wang, L., Clay, M. R., Ouma, A., Silkov, A., Liu, Y., Zhang, Z., Liu, Y., Brady, S. W., Zhou, X., Chang, T.-C., Pande, M., Davis, E., Becksfort, J., Patel, A., Wilkinson, M. R., Rahbarinia, D., Kubal, M., Maciaszek, J. L., Pastor, V., Knight, J., Gout, A. M., Wang, J., Gu, Z., Mullighan, C. G., McGee, R. B., Quinn, E. A., Nuccio, R., Mostafavi, R., Gerhardt, E. L., Taylor, L. M., Valdez, J. M., Hines-Dowell, S. J., Pappo, A. S., Robinson, G., Johnson, L.-M., Pui, C.-H., Ellison, D. W., Downing, J. R., Zhang, J. & Nichols, K. E. Genomes for Kids: The Scope of Pathogenic Mutations in Pediatric Cancer Revealed by Comprehensive DNA and RNA Sequencing. Cancer Discov. 11, 3008–3027 (2021).

71. Islam, S. M. A., Díaz-Gay, M., Wu, Y., Barnes, M., Vangara, R., Bergstrom, E. N., He, Y., Vella, M., Wang, J., Teague, J. W., Clapham, P., Moody, S., Senkin, S., Li, Y. R., Riva, L., Zhang, T., Gruber, A. J., Steele, C. D., Otlu, B., Khandekar, A., Abbasi, A., Humphreys, L., Syulyukina, N., Brady, S. W., Alexandrov, B. S., Pillay, N., Zhang, J., Adams, D. J., Martincorena, I., Wedge, D. C., Landi, M. T., Brennan, P., Stratton, M. R., Rozen, S. G. & Alexandrov, L. B. Uncovering novel mutational signatures by de novo extraction with SigProfilerExtractor. Cell Genomics 2, None (2022).

72. van der Ham, C. G., Suurenbroek, L. C., Kleisman, M. M., Antić, Ž., Lelieveld, S. H., Yeong, M., Westera, L., Sonneveld, E., Hoogerbrugge, P. M., van der Velden, V. H. J., van Leeuwen, F. N. & Kuiper, R. P. Mutational mechanisms in multiply relapsed pediatric acute lymphoblastic leukemia. Leukemia 38, 2366–2375 (2024).

73. Yan, H., Huang, J., Li, Y. & Zhao, B. Sex disparities revealed by single-cell and bulk sequencing and their impacts on the efficacy of immunotherapy in esophageal cancer. Biol. Sex Differ. 15, 22 (2024).

74. Klein, S. L. & Flanagan, K. L. Sex differences in immune responses. Nat. Rev. Immunol. 16, 626–638 (2016).

75. Goldenberg, R. L., Andrews, W. W., Faye-Petersen, O. M., Goepfert, A. R., Cliver, S. P. & Hauth, J. C. The Alabama Preterm Birth Study: intrauterine infection and placental histologic findings in preterm births of males and females less than 32 weeks. Am. J. Obstet. Gynecol. 195, 1533–1537 (2006).

76. Liu, C.-A., Wang, C.-L., Chuang, H., Ou, C.-Y., Hsu, T.-Y. & Yang, K. D. Prediction of elevated cord blood IgE levels by maternal IgE levels, and the neonate’s gender and gestational age. Chang Gung Med. J. 26, 561–569 (2003).

77. Roberts, C. W., Walker, W. & Alexander, J. Sex-Associated Hormones and Immunity to Protozoan Parasites. Clin. Microbiol. Rev. 14, 476–488 (2001).

78. Carr, B. R., Parker, C. R., Ohashi, M., MacDonald, P. C. & Simpson, E. R. Regulation of human fetal testicular secretion of testosterone: low-density lipoprotein-cholesterol and cholesterol synthesized de novo as steroid precursor. Am. J. Obstet. Gynecol. 146, 241–247 (1983).

79. Casimir, G. J., Heldenbergh, F., Hanssens, L., Mulier, S., Heinrichs, C., Lefevre, N., Désir, J., Corazza, F. & Duchateau, J. Gender differences and inflammation: an in vitro model of blood cells stimulation in prepubescent children. J. Inflamm. 7, 28 (2010).

80. Quintero, O. L., Amador-Patarroyo, M. J., Montoya-Ortiz, G., Rojas-Villarraga, A. & Anaya, J.-M. Autoimmune disease and gender: plausible mechanisms for the female predominance of autoimmunity. J. Autoimmun. 38, J109–119 (2012).

81. Moulton, V. R. Sex Hormones in Acquired Immunity and Autoimmune Disease. Front. Immunol. 9, 2279 (2018).

82. Ngo, S. T., Steyn, F. J. & McCombe, P. A. Gender differences in autoimmune disease. Front. Neuroendocrinol. 35, 347–369 (2014).

83. Wallis, C. J. D., Butaney, M., Satkunasivam, R., Freedland, S. J., Patel, S. P., Hamid, O., Pal, S. K. & Klaassen, Z. Association of Patient Sex With Efficacy of Immune Checkpoint Inhibitors and Overall Survival in Advanced Cancers: A Systematic Review and Meta-analysis. JAMA Oncol. 5, 529–536 (2019).

84. Conforti, F., Pala, L., Bagnardi, V., Pas, T. D., Martinetti, M., Viale, G., Gelber, R. D. & Goldhirsch, A. Cancer immunotherapy efficacy and patients’ sex: a systematic review and meta-analysis. Lancet Oncol. 19, 737–746 (2018).

85. Milne, E., Greenop, K. R., Scott, R. J., Bailey, H. D., Attia, J., Dalla-Pozza, L., de Klerk, N. H. & Armstrong, B. K. Parental prenatal smoking and risk of childhood acute lymphoblastic leukemia. Am. J. Epidemiol. 175, 43–53 (2012).

86. Chang, J. S., Selvin, S., Metayer, C., Crouse, V., Golembesky, A. & Buffler, P. A. Parental smoking and the risk of childhood leukemia. Am. J. Epidemiol. 163, 1091–1100 (2006).

87. Chunxia, D., Meifang, W., Jianhua, Z., Ruijuan, Z., Xiue, L., Zhuanzhen, Z. & Linhua, Y. Tobacco smoke exposure and the risk of childhood acute lymphoblastic leukemia and acute myeloid leukemia: A meta-analysis. Medicine (Baltimore) 98, e16454 (2019).

88. Metayer, C., Zhang, L., Wiemels, J. L., Bartley, K., Schiffman, J., Ma, X., Aldrich, M. C., Chang, J. S., Selvin, S., Fu, C. H., Ducore, J., Smith, M. T. & Buffler, P. A. Tobacco Smoke Exposure and the Risk of Childhood Acute Lymphoblastic and Myeloid Leukemias by Cytogenetic Subtype. Cancer Epidemiol. Biomark. Prev. Publ. Am. Assoc. Cancer Res. Cosponsored Am. Soc. Prev. Oncol. 22, 1600–1611 (2013).

89. Xu, K., Li, S., Whitehead, T. P., Pandey, P., Kang, A. Y., Morimoto, L. M., Kogan, S. C., Metayer, C., Wiemels, J. L. & de Smith, A. J. Epigenetic Biomarkers of Prenatal Tobacco Smoke Exposure Are Associated with Gene Deletions in Childhood Acute Lymphoblastic Leukemia. Cancer Epidemiol. Biomark. Prev. Publ. Am. Assoc. Cancer Res. Cosponsored Am. Soc. Prev. Oncol. 30, 1517–1525 (2021).

90. de Smith, A. J., Kaur, M., Gonseth, S., Endicott, A., Selvin, S., Zhang, L., Roy, R., Shao, X., Hansen, H. M., Kang, A. Y., Walsh, K. M., Dahl, G. V., McKean-Cowdin, R., Metayer, C. & Wiemels, J. L. Correlates of Prenatal and Early-Life Tobacco Smoke Exposure and Frequency of Common Gene Deletions in Childhood Acute Lymphoblastic Leukemia. Cancer Res. 77, 1674–1683 (2017).

91. Liu, T., Xu, K., Pardeshi, A., Myint, S. S., Kang, A. Y., Morimoto, L. M., Lieber, M. R., Wiemels, J. L., Kogan, S. C., Metayer, C. & de Smith, A. J. Early-life tobacco exposure is causally implicated in aberrant RAG-mediated recombination in childhood acute lymphoblastic leukemia. Leukemia 38, 2492–2496 (2024).

92. Gao, Q., Ryan, S. L., Iacobucci, I., Ghate, P. S., Cranston, R. E., Schwab, C., Elsayed, A. H., Shi, L., Pounds, S., Lei, S., Baviskar, P., Pei, D., Cheng, C., Bashton, M., Sinclair, P., Bentley, D. R., Ross, M. T., Kingsbury, Z., James, T., Roberts, K. G., Devidas, M., Fan, Y., Chen, W., Chang, T.-C., Wu, G., Carroll, A., Heerema, N., Valentine, V., Valentine, M., Yang, W., Yang, J. J., Moorman, A. V., Harrison, C. J. & Mullighan, C. G. The genomic landscape of acute lymphoblastic leukemia with intrachromosomal amplification of chromosome 21. Blood 142, 711–723 (2023).

93. Nielsen, K. P., Zhao, L., Stamnes, J. J., Stamnes, K. & Moan, J. The importance of the depth distribution of melanin in skin for DNA protection and other photobiological processes. J. Photochem. Photobiol. B 82, 194–198 (2006).

94. Jones, C. L., Degasperi, A., Grandi, V., Amarante, T. D., Genomics England Research Consortium, Mitchell, T. J., Nik-Zainal, S. & Whittaker, S. J. Spectrum of mutational signatures in T-cell lymphoma reveals a key role for UV radiation in cutaneous T-cell lymphoma. Sci. Rep. 11, 3962 (2021).

95. Maia, A. T., Tussiwand, R., Cazzaniga, G., Rebulla, P., Colman, S., Biondi, A. & Greaves,M. Identification of preleukemic precursors of hyperdiploid acute lymphoblastic leukemia in cord blood. Genes. Chromosomes Cancer 40, 38–43 (2004).

96. Maia, A. T., van der Velden, V. H. J., Harrison, C. J., Szczepanski, T., Williams, M. D., Griffiths, M. J., van Dongen, J. J. M. & Greaves, M. F. Prenatal origin of hyperdiploid acute lymphoblastic leukemia in identical twins. Leukemia 17, 2202–2206 (2003).

97. de Smith, A. J., Wahlster, L., Jeon, S., Kachuri, L., Black, S., Langie, J., Cato, L. D., Nakatsuka, N., Chan, T.-F., Xia, G., Mazumder, S., Yang, W., Gazal, S., Eng, C., Hu, D., Burchard, E. G., Ziv, E., Metayer, C., Mancuso, N., Yang, J. J., Ma, X., Wiemels, J. L., Yu, F., Chiang, C. W. K. & Sankaran, V. G. A noncoding regulatory variant in IKZF1 increases acute lymphoblastic leukemia risk in Hispanic/Latino children. Cell Genomics 4, 100526 (2024).

98. Martin, A. R., Kanai, M., Kamatani, Y., Okada, Y., Neale, B. M. & Daly, M. J. Current clinical use of polygenic scores will risk exacerbating health disparities. Nat. Genet. 51, 584–591 (2019).

99. Jeon, S., Lo, Y. C., Morimoto, L. M., Metayer, C., Ma, X., Wiemels, J. L., de Smith, A. J. & Chiang, C. W. K. Evaluating genomic polygenic risk scores for childhood acute lymphoblastic leukemia in Latinos. Hum. Genet. Genomics Adv. 4, 100239 (2023).

100. Im, C., Raduski, A., Mills, L. J., Johnson, R. A., DeWan, A., Ma, X., Wiemels, J. L., Metayer, C., Yang, J. J., Nelson, H. H., Yang, T., Basu, S., Turcotte, L. M., Pankratz, N., Scheurer, M. E. & Spector, L. G. Abstract 2845: Novel genetic risk loci for B-cell acute lymphoblastic leukemia in African American children: Findings from the Admixture and Risk of Acute Leukemia (ADMIRAL) Study. Cancer Res. 84, 2845 (2024).

101. Brady, S. W., Liu, Y., Ma, X., Gout, A. M., Hagiwara, K., Zhou, X., Wang, J., Macias, M., Chen, X., Easton, J., Mulder, H. L., Rusch, M., Wang, L., Nakitandwe, J., Lei, S., Davis, E. M., Naranjo, A., Cheng, C., Maris, J. M., Downing, J. R., Cheung, N.-K. V., Hogarty, M. D., Dyer, M. A. & Zhang, J. Pan-neuroblastoma analysis reveals age- and signature-associated driver alterations. Nat. Commun. 11, 5183 (2020).

